# A row-generation approach for simultaneous multiple needle trajectory planning in radiofrequency ablation

**DOI:** 10.1101/2021.06.11.21258244

**Authors:** Shefali Kulkarni-Thaker, Dionne Aleman, Aaron Fenster

## Abstract

Radiofrequency ablation is a thermal therapy for moderately-sized cancerous tumors. A target is killed with high temperatures obtained due to the current passed through one or more electrodes (needles) inserted into it. The needles’ trajectory must be meticulously planned to prevent interference with dense organs like bone or puncturing of critical structures like veins. By approximating the thermal lesion to an ellipse, we predefine several valid needle trajectories and then solve an integer programming model to identify pairwise valid needle positions, that meet clinical criteria, using a variation of the classic set cover model. To improve the models’ tractability and scalability, we use row generation-based decomposition techniques that determines pairwise validity using two different types of cuts. Finally, we analyze target and organ-at-risk (OAR) damage using several thermal damage models. Our method is tested on 12 liver targets: three targets each with four different surgical margins. We show promising results that meet clinical guidelines while obtaining full target coverage.

## 1 Introduction

Radiofrequency ablation (RFA) is a minimally invasive cancer treatment that destroys cancerous tumours (targets) by exposing them to very high heat. The heat is delivered through electrodes (called needles) that are inserted directly into the targets, either percutaneously, laproscopically, or via open surgery. Because heat is only delivered at the tip of the needle, fewer healthy tissues are exposed and there are fewer side effects than in radiation therapy, where all tissues along the paths of the beams are exposed. However, inaccurate placement of the needles is common, resulting in failure to eradicate cancerous tissue or excessive damage to surrounding healthy tissue, and thus CT image guidance is used to help accurately position the needles, although at the cost of radiation exposure that may render CT usage—and therefore highly conformal treatments—unacceptable for some patients. New ultrasound guidance techniques achieve similar accuracy without radiation, and therefore allow for nuanced treatments to be planned for any patient [1]. While mathematical methods to optimize treatment plans have been successful in radiation therapy treatment modalities (e.g., Intensity modulated radiation therapy, stereotactic radiosurgery, brachytherapy), there are few similar attempts for RFA, and most consider only the impact of the needle on the targets, as opposed to the full needle trajectory, which may render many potential solutions undeliverable (e.g., if the needle must pass through bone or blood vessels to reach the desired position). We therefore propose a mathematical optimization approach to design RFA treatments with consideration of needle trajectories.

There are several kinds of ablation needles. For instance, an ablation needle can be clustered, an equipment with multiple equidistant tines, or it can be umbrella shaped protracted needle. Most commonly used ablation needle is a single needle or set of single needles called as multiple needles, and is the focus of this work. Ablation needles are up to 30 cm long, and have conducting and insulating parts (Figure 1). Typically, a needle is inserted so that the entire conducting part (up to 4 cm long) is within the target. An RFA devices transmits electricity through the needle which is converted to heat due to frictional resistance. Since, only the conducting portion placed within target is able to transmit heat, the thermal spread is controlled and localized making RFA a minimally invasive treatment option for moderately-sized superficial tumors.

**Figure 1:**
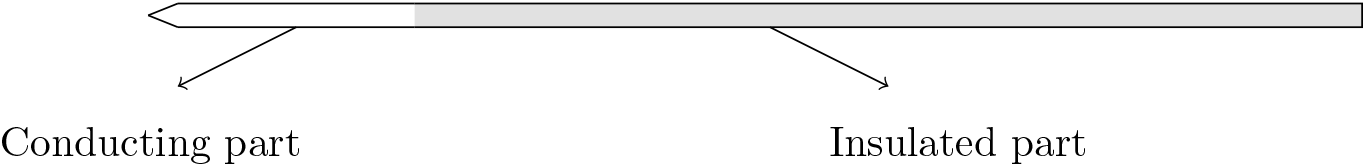
A typical single ablation needle

Inverse treatment planning for RFA identifies the needle’s position and orientation to determine optimal thermal dose delivery. A 3D box around the liver target is divided in to unit grids called voxels (“volume pixels”), where each voxel as a target, an organ-at-risk (OAR), or a forbidden path voxel (e.g., a blood vessel or rib). Algorithms for needle placement and thermal dose delivery that achieve full target damage are then designed. Needle placements must consider: (1) physician and patient positions as well as its angle of entry to avoid needle gliding, (2) presence of critical organs along the trajectory, and (3) the intersection of conducting tines with OAR. Finally, after needle placement, thermal dose delivery must consider appropriate treatment voltage or power for full target coverage while avoiding unnecessary OAR damage.

Thermal dose delivery must be computed using simultaneous partial differential equations (PDE), Laplacian, the electrostatic equation, and the Pennes’ bioheat transfer equation (BHTE) [2]. BHTE computes the temperature of a voxel at a given time while Laplacian computes the power absorbed from the source. Although BHTE accounts for voxel temperature, it does not consider voxel’s length of exposure at given temperature. This thermal history of a voxel provides insight into cellular damage and is computed using the non-linear Arrhenius thermal damage model (ATDM) [3–6] that uses input from BHTE to determine percentage of cellular damage. The computational expense of solving PDEs and non-linearity of thermal dose models, makes the delivery of inverse plans computationally as well as mathematically challenging.

Previous RFA works are categorized based on their methodology to determine thermal dose. Inexact methods use unconstrained linear models to determine needle position where the voxels enclosed within a thermal lesion, with a pre-defined radius, are considered ablated [7–10]. Exact methods [11–14] determine needle position and orientation by solving a nonlinear optimization model, with fixed treatment times, that are constrained by BHTE and Laplacian equations. Thus, at every new position and orientation required recomputation of the PDEs. Incorporating multi-needle ablations and trajectory planning can be difficult with PDE-constrained systems, though we have previously prposed optimization approaches and solution methodologies to solve such problems for RFA treatment design [15].

Trajectory planning has been explored in the literature where a list of acceptable trajectories is typically proposed using heuristics [16–22] and the final selection is performed using computer-assisted visualization where each path is rated based on a linear combination of several criteria or a Pareto-optimal front [16, 19]. Pareto-optimal fronts have been explored in a set cover formulation [23, 24] while considering the possibility of needle pull-back, though not needle trajectory; we note that these particular publications borrow from our earlier RFA set cover formulations [25]. **(author?)** [12] used convex functions to describe forbidden regions and developed semi-infinite techniques for single needle trajectory planning. While most works focus on single needle trajectory planning, heuristics for sequential placement [18] and integer models for simultaneous placement [21, 22] of multiple needles have been explored. Sequential techniques use integer models to first identify a minimum number of trajectories required for target coverage followed by a minimum number of ablations required on those trajectories, resulting in inherently suboptimal solutions [21, 22]. Path length, angle of entry, and proximity to critical structures are used to determine acceptable trajectories.

Previously, we approached the RFA inverse planning problem in two stages [15]. In the first stage, referred to as needle orientation optimization (NOO), geometric shape approximations are used to compute the needle’s position and orientation for single, multiple, or clustered needles. In the second stage, called as thermal dose optimization (TDO), NOO solution is used to compute the Laplacian equation with constant electrical conductivity to determine the specific absorption rate for each voxel. The results from Laplacian are used to compute the BHTE which in turn determines the cellular damage using ATDM. Using this information we determine thermal damage incurred using various damage models: threshold temperature, Arrhenius damage index, and percentage of cellular damage. Using the same framework, we now incorporate trajectory planning for multiple needle placement in the NOO stage.

A set of valid ellipses corresponding to valid needle trajectories and ablation centers are determined through heuristics. Then, a variation of the classic set cover-based integer model is used to identify a union of ellipses that given full target coverage and minimal OAR coverage. This variation allows for selection of multiple ellipses that satisfy clinical needle placement criteria for each needle pair, called as pairwise validity. Although set cover has been proposed for needle placement as previously discussed, consideration of pairwise validity, as well as our cut-based optimization approach, is novel.

To improve the tractability, we use row generation-based decomposition techniques, where the reduced master problem ensures full target coverage and the feasibility check subproblem ensures pairwise validities. For pairwise validity, we generate and compare two types of cuts: pairwise no-good and group cuts. Finally, we perform TDO to determine target and OAR thermal damage. Thus, a treatment plan, where needle trajectories, number of needles with their centers and orientations, and source voltage for 100% target kill, is determined. This approach provides the flexibility to use needles with heterogeneous tip lengths along with non-parallel needle placement.

## 2 Trajectory planning

Coverage of a voxel by an ellipse can be determined given a needle position and its vendor-specified thermal lesion radius. We predefine all ellipses that cover target voxels, and ellipses produced due to invalid trajectories are rejected. A trajectory can be invalid due to its intersection with critical structures like bone or vessels, due to physician discomfort (e.g., a physician will avoid inserting needle from below a supine patient), or due to an insertion angle that may cause the needle to slide.

Clinically, needles are inserted sequentially, but are not necessarily removed sequentially. Thus, needle paths cannot cross. Further, even if each needle is removed before the next needle is inserted, it is undesirable to place a needle center in ablated tissue as the ablated tissue will prevent the heat from transferring to non-ablated cells due to thermal equilibrium, and therefore needle paths still cannot cross even in a fully sequential insertion process. Additionally, vendor specifications recommend a minimum distance between the needle centers to achieve a target thermal lesion, which is obtained clinically using a separator which enforces parallel needle placement (which we consider to be a single clustered needle scenario), preventing the treatment flexibility that could be gained in a multiple needle scenario; however, this minimum distance can be enforced through constraints in an optimization approach to needle placement. Finally, clinical studies indicate poor ablation for large inter-needle angles [26, 27]. We therefore develop pairwise needle orientation constraints to ensure that (1) the conducting portion of the needles do not intersect, (2) the distance between two needle centers is ≥ *ω*, and (3) the angle between two needles’ major axes is ≤ *α*. By ensuring that each pair of needles satisfies these rules, all needles satisfy the rules.

The IP approaches presented here (Section 2.2) require creation of all the valid trajectories (i.e., ellipses) a priori, which can have significant overhead, especially when high accuracy in needle placement is desirable. However, ellipse creation (Section 2.1) is an embarrassingly parallel problem and runtimes can be significantly improved. Further, the trajectory planning models assume a vendor-specified lesion volume for each ellipse or needle, although, in practice, thermal doses will vary from this estimation due to several factors including local tissue properties and interactions. Additionally, a target or OAR voxel may be covered multiple times due to overlapping ellipses but the actual dose at a voxel is not additive. Therefore, during the TDO stage (Section 3), we disregard the assumption on lesion sizes and compute true thermal distributions.

### 2.1 Ellipse definition

Recall that a single needle can be defined as an ellipse with the ellipse center at the center of the needle’s conducting tip and the ellipse size and shape (including the principal axis, which is the needle itself) determined by vendor-provided ablation specifications. To generate a set of candidate ellipses, we consider every non-boundary target voxel to be a potential ellipse center, and for each center, the set of all valid ellipse orientations (principal axis vectors) are potential orientations. Orientations are obtained by enumerating all position vectors between the target’s geometric center voxel and the boundary voxels:

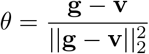

where **v** is the 3D co-ordinate of a target voxel and **g** is the target centroid given by

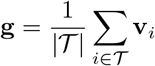

A valid orientation adheres to user-specified rules, e.g., entry from the top of the patient and non-oblique entry at the patient surface; while valid orientations at an ellipse center are those that do not cause intersection between the needle conducting tip and forbidden voxels (OARs, bones, and veins). Since this process results in an extremely large number of ellipses, for computational tractability, we sample the full set of candidate ellipses by only considering every *p*th ellipse center and only *n* orientations. The *n* orientations are selected uniformly from the orientations that create angles in the range [30^°^, 150^°^] with the patient surface (where 90^°^ is orthogonal to the patient surface), since larger insertion angles correspond to easier clinical delivery. Both *p* and *n* are user-provided. Based on experimentation for tractability, we use *n* = 20 and

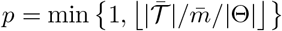

where 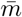 is the user-provided maximum number of ellipses desired, 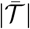 is the cardinality of all the valid centers, and |Θ| = *n*, where Θ is the set of all orientations. Algorithm 1 shows the steps to define ellipsoids with valid trajectories.

#### Algorithm 1

Define ellipse set

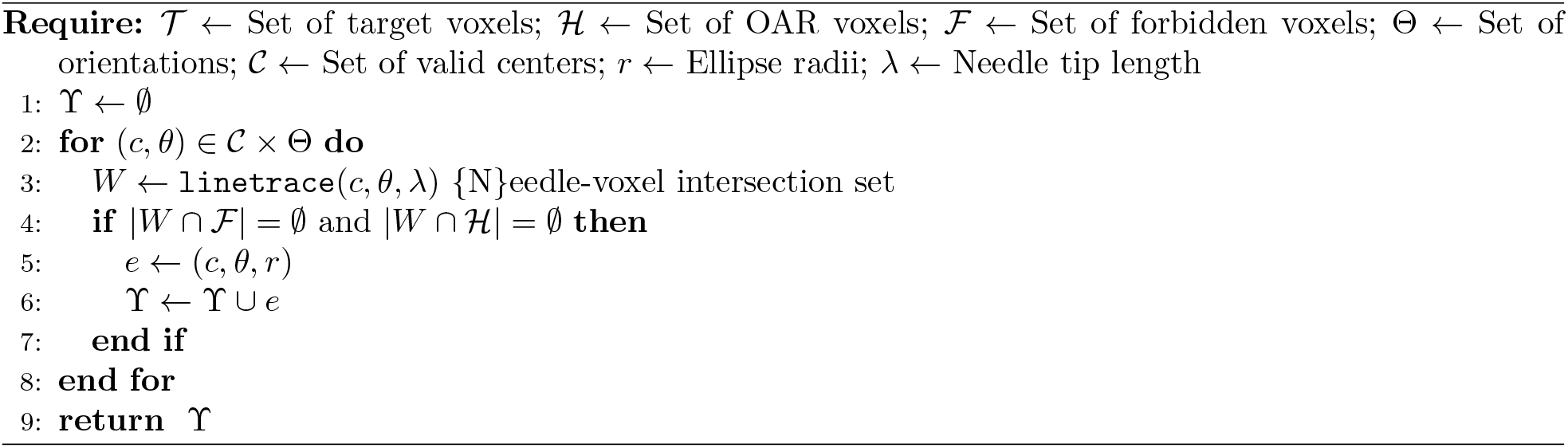

For each ellipse *e* = (**c, *θ*, r**), its target and OAR coverage is determined by translating and rotating the target and OAR voxels around the center **c** and orientation ***θ***. A voxel *v* ∈ 𝒯 ∪ ℋ is covered by an ellipse with radii **r** centered at the origin and parallel to the coordinate axes if

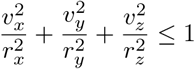

Finally, we create an incidence matrix **E** of size |𝒯 | × |𝓇| to determine if target voxel **v** ∈ 𝒯 is covered by ellipse *e*_*i*_ ∈ 𝓇, and the cost *f*_*i*_ of selecting ellipse *e*_*i*_ the total OAR voxels it covers.

### 2.2 Integer model

Let *ℱ* and 𝓇 be the set of forbidden structures and predefined valid ellipses, respectively. The goal is to cover each target voxel at least once, with at least *<u>k</u>* and at most 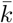 ellipses or sets, while adhering to pairwise validity constraints. This formulation is a variation of the classic set cover problem (SCP) model, and is given by

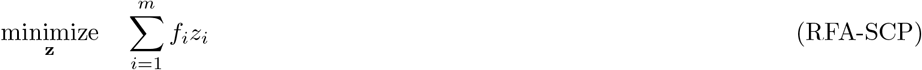

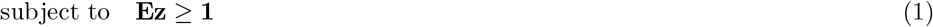

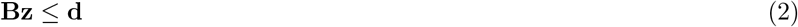

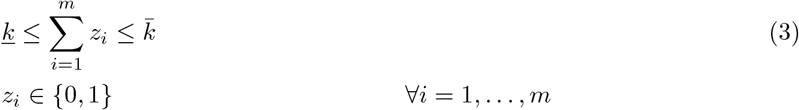

where *m* = |𝓇| is the number of predefined ellipses, *z*_*i*_ indicates if ellipse *e*_*i*_ is chosen, **E** is a |𝒯 |×|𝓇| incidence matrix that indicates if target voxel *j* ∈ 𝒯 is covered by ellipse *e*_*i*_, each row of matrix **B** identifies one or more invalid ellipse pairs, and *f*_*i*_ is the cost of ellipse *e*_*i*_ defined by its total OAR voxel coverage. Thus, the objective function determines the total cost of selected sets (ellipses), thereby minimizing OAR coverage. Constraint 1 ensures that each target voxel is covered at least once by the union of selected ellipses. Due to localized nature of RFA treatment, coverage of every single voxels is essential as needle placements can fail to cover either internal or boundary target voxels due to insufficient heat deposition. Constraint 2 eliminates selection of invalid ellipse pairs and Constraint 3 bounds the minimum and maximum number of ellipses or ablations.

The pairwise validity matrix, **B**, consists of either pairwise no-good or group cuts. A no-good cut is an inequality that enforces at least one binary variable to change its value. For an ellipse pair (*e*_*i*_, *e*_*p*_), a pairwise no-good cut is given by

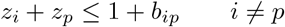

where *b*_*ip*_ ∈ {0, 1} indicates if ellipses *e*_*i*_ and *e*_*p*_ are pairwise valid. If ellipse pair (*e*_*i*_, *e*_*p*_) is invalid, then *b*_*ip*_ = 0, which enforces selection of either *e*_*i*_ or *e*_*p*_ but not both. Thus, each pairwise cut is a no-good cut that eliminates a single ellipse pair and if all the ellipse pairs were invalid, indicating either a single ablation or an infeasible multiple needle solution, it would generate *m*!*/*2!(*m*— 2)! constraints. For *m*≈ 4, 000, there are up to 3,000,000 no-good cuts (Case 3M).

To reduce the number of pairwise validity constraints and improve tractability, we propose a variation to pairwise cuts where for each ellipse *e*_*i*_, we generate a single group cut of the form

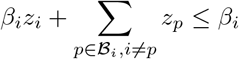

where ℬ_*i*_ is the set of ellipses that form invalid pairs with ellipse *e*_*i*_ and |*ℬ*_*i*_| = *β*_*i*_. If *β*_*i*_ = *m* − 1, then ellipse *e*_*i*_ does not form a valid pair with any other ellipses. Thus, unlike pairwise cuts, each group cut eliminates *β*_*i*_ ellipse pairs. Further, at most *m* cuts are required to eliminate all invalid ellipse pairs, significantly improving tractability over pairwise no-good cuts.

The model RFA-SCP requires a priori creation of the target coverage (**E**) and pairwise validity (**B**) matrices, potentially resulting in a memory intensive model that may not scale to large targets. To improve computational runtime as well as to overcome memory limitations, we design a decomposition technique based on row generation. We first solve the model RFA-SCP with only a subset of constraints, called the reduced master problem (RMP). Violated constraints, obtained through a feasibility check (FC), are added to the RMP which is then resolved. The process is continued until all constraints are satisfied. We explore a row generation approach on target coverage as well as on pairwise validity.

#### 2.2.1 Target coverage row generation

The RMP when performing row generation on target coverage is given by:

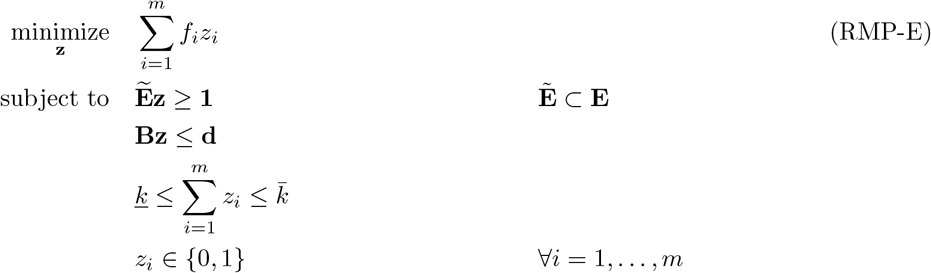

For the first iteration, we consider only boundary target voxels (i.e.,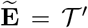), obtained by the grassfire algorithm. We hope that by covering the boundary of the target, we also cover the target interior. However, it is possible that the union of selected ellipses does not cover a subset of central target voxels. Using the RMP-E solution 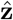, we perform a feasibility check to ensure that all target voxels are indeed covered:

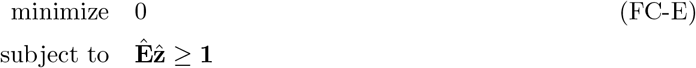

Each violated inequality 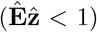 indicates an uncovered target voxel and all such violations are added to the matrix 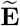 in model RMP-E, which is resolved. The process is continued until FC-E returns no cuts, at which point full target coverage is achieved. If group cuts are used to determine the pairwise validity matrix, then **B** ∈ *ℛ* ^*m*×*m*^. Further, the boundary of the target can be up to 50% of the entire target, which significantly decreases the number of the constraints, making the model less memory intensive and therefore more tractable. However, this approach requires the time consuming a priori creation of the pairwise validity matrix.

### 2.2.2 Pairwise validity row generation

The RMP is given by:

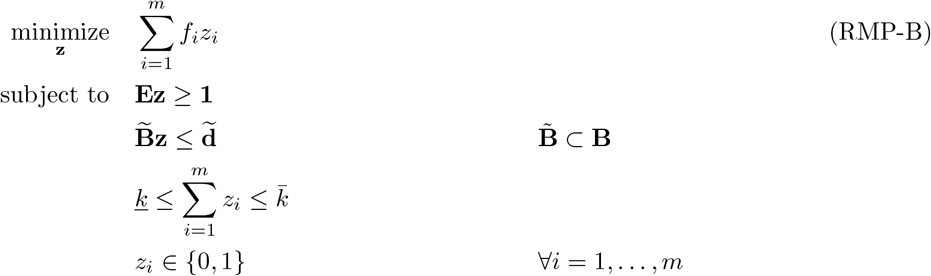

where 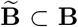 and 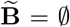 for the first iteration. The selected ellipses, 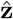, are passed to the feasibility check subproblem:

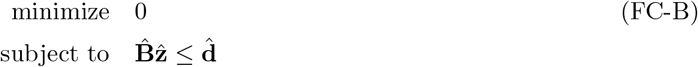

where 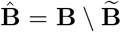. The FC-B is solved algorithmically to identify invalid pairs instead of simply indicating the presence of invalid pairs, eliminating a priori creation of **B**. Pairwise cuts (Algorithm 2) or group cuts (Algorithm 3) are generated using a set of rules and cuts are passed to the RMP-B.

#### Algorithm 2

Create pairwise no-good cuts

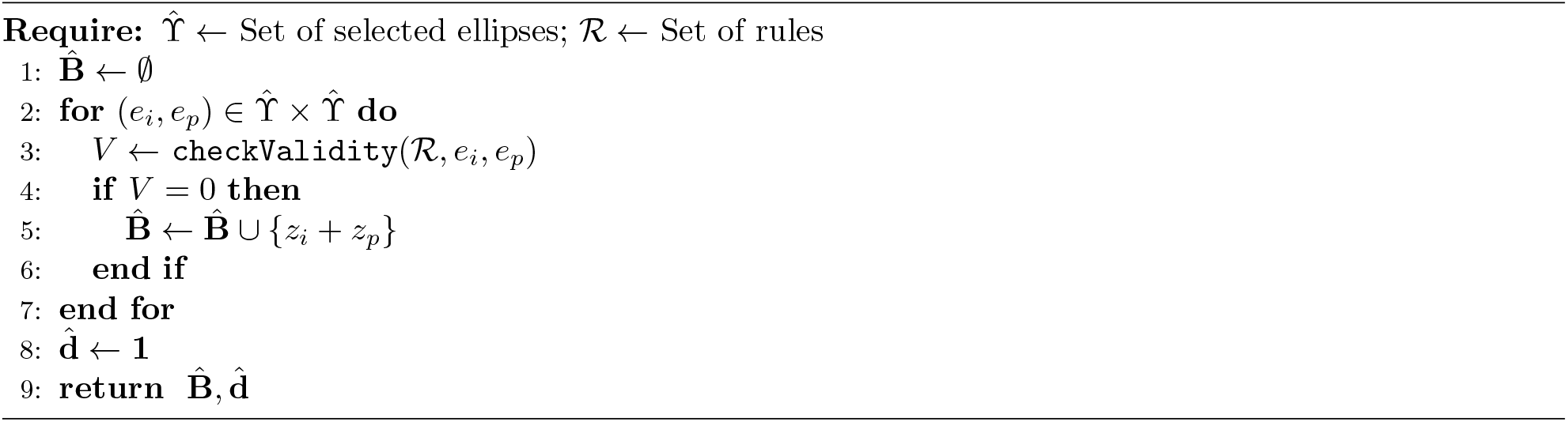

#### Algorithm 3

Create group cuts

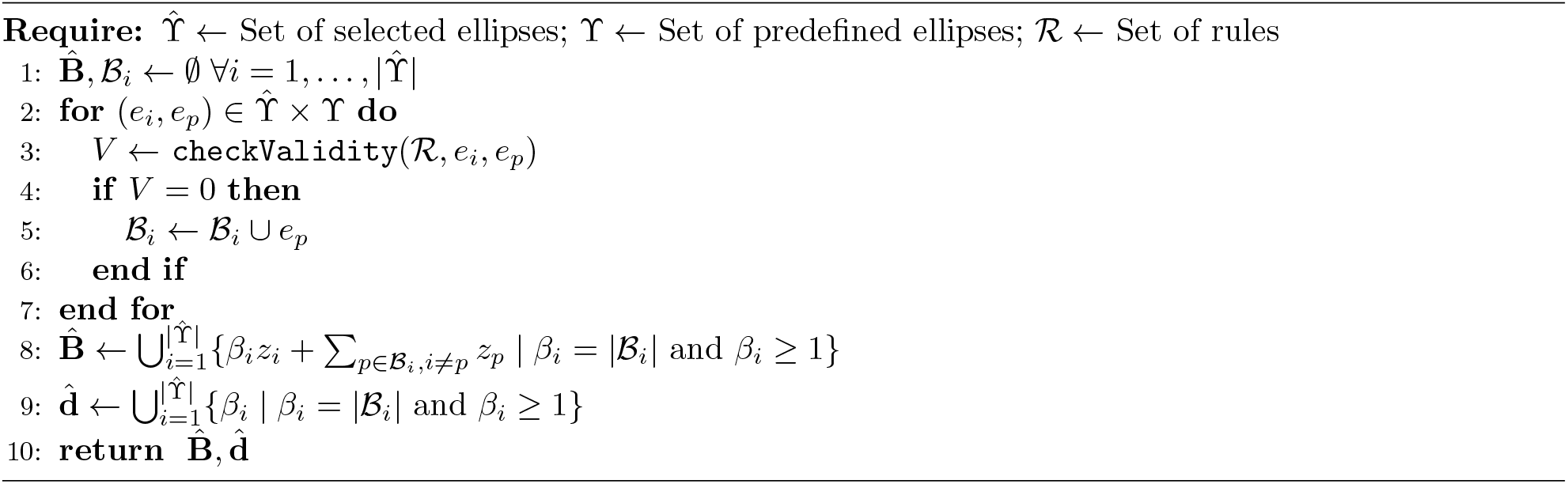

#### Algorithm 4

checkValidity: Pairwise validity check

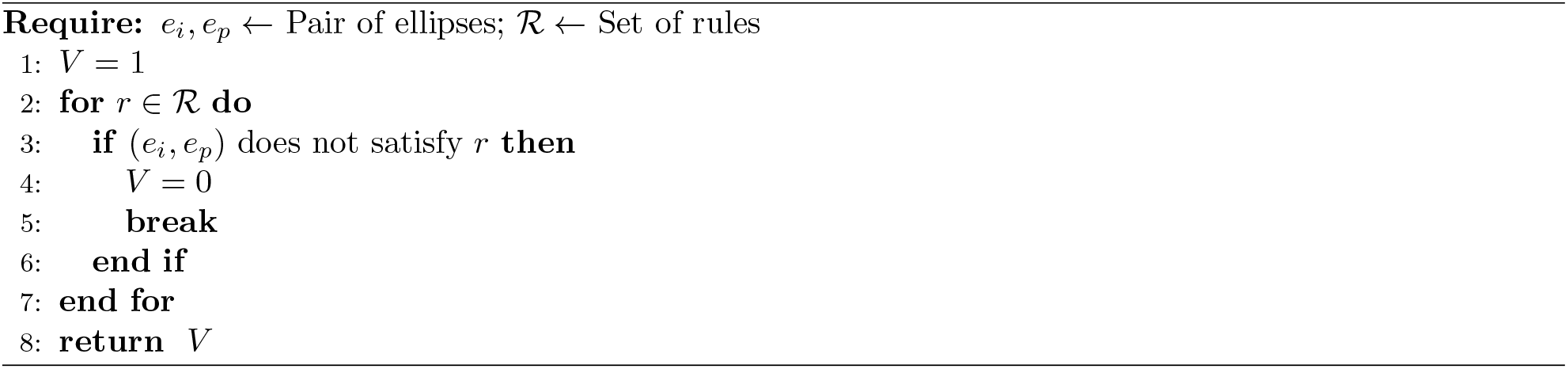

To account for clinical practices, we solve model RFA-SCP with unbounded 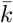 or bounded by a small finite number to account for patient discomfort since a high number of ablations is not desired. If a fixed set of *k* needles must be used, then we set 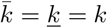. Depending on the maximum ellipse coverage, the value of <u>*k*</u> is selected as follows:

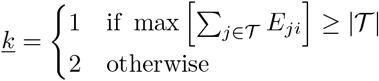

## 3 Thermal dose optimization

Up on identification of needle positions and orientations, we compute thermal dose thereby lifting the geometric assumption on the shape of thermal lesion made in NOO. Thermal dose is determined by solving BHTE and ATDM. In a 3D system, BHTE is given by [2, 28]

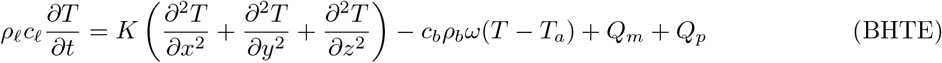

where *ρ*_*ℓ*_ and *ρ*_*b*_ are the densities of tissue and blood (kg/m^3^), respectively; *c*_*ℓ*_ and *c*_*b*_ are the specific heats of the tisue and blood (J/kg-K), respectively; *K* is the thermal conductivity of the tissue (W/m-K); *ω* is the blood perfusion coefficient, i.e., blood flow rate/unit mass tissue (1/s); *T* and *T*_*a*_ are the temperatures of tissue and arterial blood (K), respectively; *Q*_*p*_ is the power absorbed per unit volume of the tissue (W/m^3^); and *Q*_*m*_ is metabolic heating, which is usually considered negligible [29]. The heat source, *Q*_*p*_, is approximated by [13, 14]

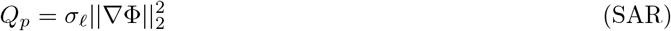

where Φ and *σ*_*ℓ*_ are the electric potential and tissue electrical conductivity, respectively. Assuming a constant electrical conductivity, the electric potential is given by the Laplacian [30] as follows:

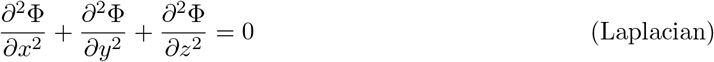

The needle is positioned so that the center of its conducting part is placed at the ellipse enter obtained from NOO and a ray tracing algorithm is used to compute the needle-voxel intersection set [31]. For Laplacian, the initial conditions (voltage) are set to 0 for all voxels except the needle-voxel intersection set, whose initial conditions are set to the input voltage of the needle. Both BHTE and Laplacian are solved using a finite difference scheme with Dirichlet boundary conditions.

ATDM [3–6] considers thermal history of a voxel, i.e., how long a voxel is exposed to a given temperature, and computes cumulative thermal damage over a period of time. ATDM is a dimensionless number Ω_*js*_ computed for every voxel *j* as follows:

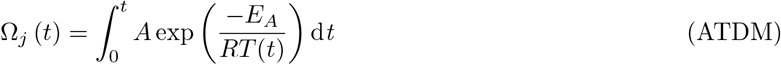

where *A* is the frequency factor, *E*_*A*_ is the activation energy, and *R* is the universal gas constant. *T* (*t*) is the average tissue absolute temperature (i.e., temperature in Kelvin) in the time interval [0, *t*] obtained from BHTE. Physically, Ω_*j*_ is described as [32]

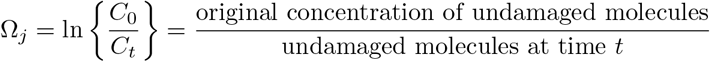

Thus, if *C*_*t*_ ∈ [0, 1] and *C*_0_ = 1, then percentage of damaged molecules at time *t* is

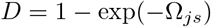

because exp(− Ω_*js*_) = *C*_*t*_ = undamaged molecules at time *t*. We describe these percentage damage models as D63 for *p* = 63% tissue damage, D70 for *p* = 70% tissue damage, etc. A value of *p* = 0.63 or 63% is associated with irreversible thermal damage and corresponds to Ω_*j*_ = 1.

We define a set of needle configurations as a combination of single needle lengths (*λ* ∈ Λ) and source voltage (*ϕ* ∈ 𝒱). The set of damage models is given by: *d* ∈ *𝒟* = {BHTE, ATDM, D63, D70, D80, D95}. For each needle configuration, (*ϕ, λ*), we first compute the BHTE for fixed treatment time using inputs from Laplacian and then compute the ATDM followed by the percentage damage models. Finally, we determine the minimum treatment voltage and treatment times for a full target coverage [15].

## 4 Results

We perform experiments on three clinical liver cases (Robarts Research Institute, Western University) to which we add surgical margins of 0 mm (N), 3 mm (S), 5 mm (M), and 10 mm (L) around the target to ensure microscopic tumor particle coverage (Table 1). Computations are conducted on AMD Opteron^™^4332 HE CPU with 3 GHz using MATLAB R2015b (Mathworks, Inc.).

**Table 1:**
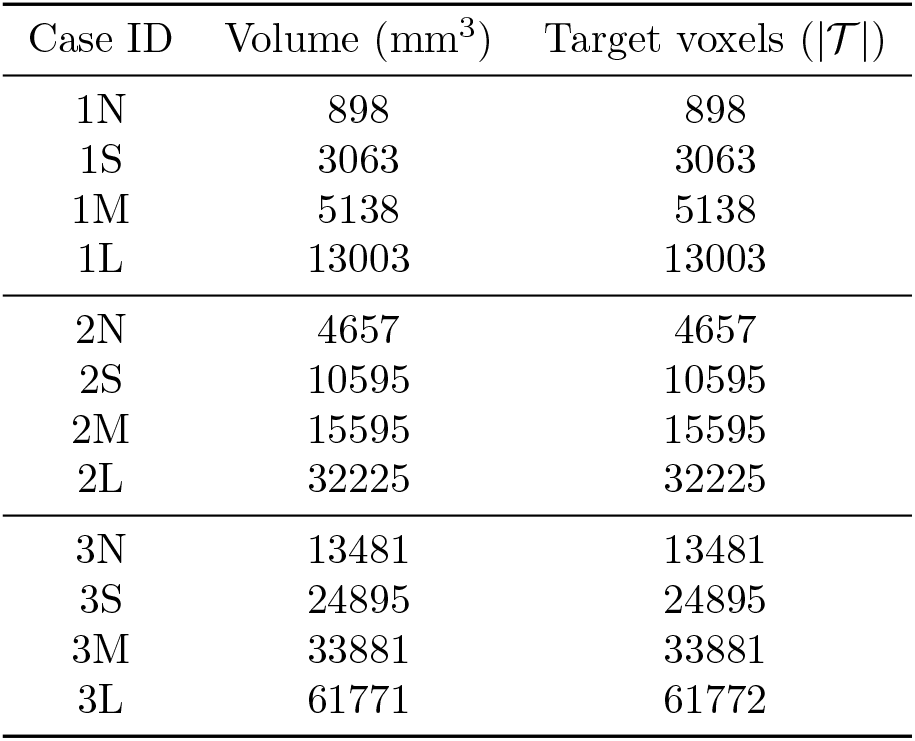
Description of case studies

| Case ID | Volume (mm<sup>3</sup>) | Target voxels ($\vert \mathcal{T}\vert $) |
| --- | --- | --- |
| 1N | 898 | 898 |
| 1S | 3063 | 3063 |
| 1M | 5138 | 5138 |
| 1L | 13003 | 13003 |
| 2N | 4657 | 4657 |
| 2S | 10595 | 10595 |
| 2M | 15595 | 15595 |
| 2L | 32225 | 32225 |
| 3N | 13481 | 13481 |
| 3S | 24895 | 24895 |
| 3M | 33881 | 33881 |
| 3L | 61771 | 61772 |

We consider five needle tip lengths (mm), Λ = {7, 10, 20, 30, 40}. For each of the 12 cases, we generate ellipses for each tip length, resulting in a maximum of 60 runs (12 cases ×5 needle tips). We control the number of ellipses generated with an upper bound. Therefore, while the maximum number of ellipses generated is |*𝒞*|× |Θ|, the actual number of ellipses generated is less due to elimination of invalid ellipses (Table 2). We note that certain combinations, e.g., Case 1N and *λ* ≥ 20 mm or Case 2N and *λ* ≥ 40 mm, do not produce any valid ellipses because the needle tip length is longer than the target size resulting in intersection with the OAR voxels. Thus, although in general increasing numbers of ellipses increases runtime, the actual runtime is affected by preprocessing to reject invalid ellipses and paths (Figure 2(a)).

**Table 2:** Total ellipses generated

| Case ID | Tip length ($\lambda$ mm) | | | | |
| --- | --- | --- | --- | --- | --- |
|  | 7 | 10 | 20 | 30 | 40 |
| 1N | 396 | 169 | - | - | - |
| 1S | 831 | 437 | 37 | - | - |
| 1M | 1182 | 775 | 72 | - | - |
| 1L | 1572 | 1246 | 228 | 4 | - |
| 2N | 1599 | 986 | 254 | 7 | - |
| 2S | 2211 | 1478 | 440 | 61 | - |
| 2M | 2606 | 2037 | 783 | 128 | - |
| 2L | 2696 | 2270 | 1147 | 339 | 33 |
| 3N | 2780 | 1927 | 979 | 356 | 22 |
| 3S | 3649 | 2735 | 1236 | 442 | 76 |
| 3M | 3740 | 3001 | 1577 | 642 | 168 |
| 3L | 4430 | 3712 | 2271 | 953 | 291 |

**Figure 2:**
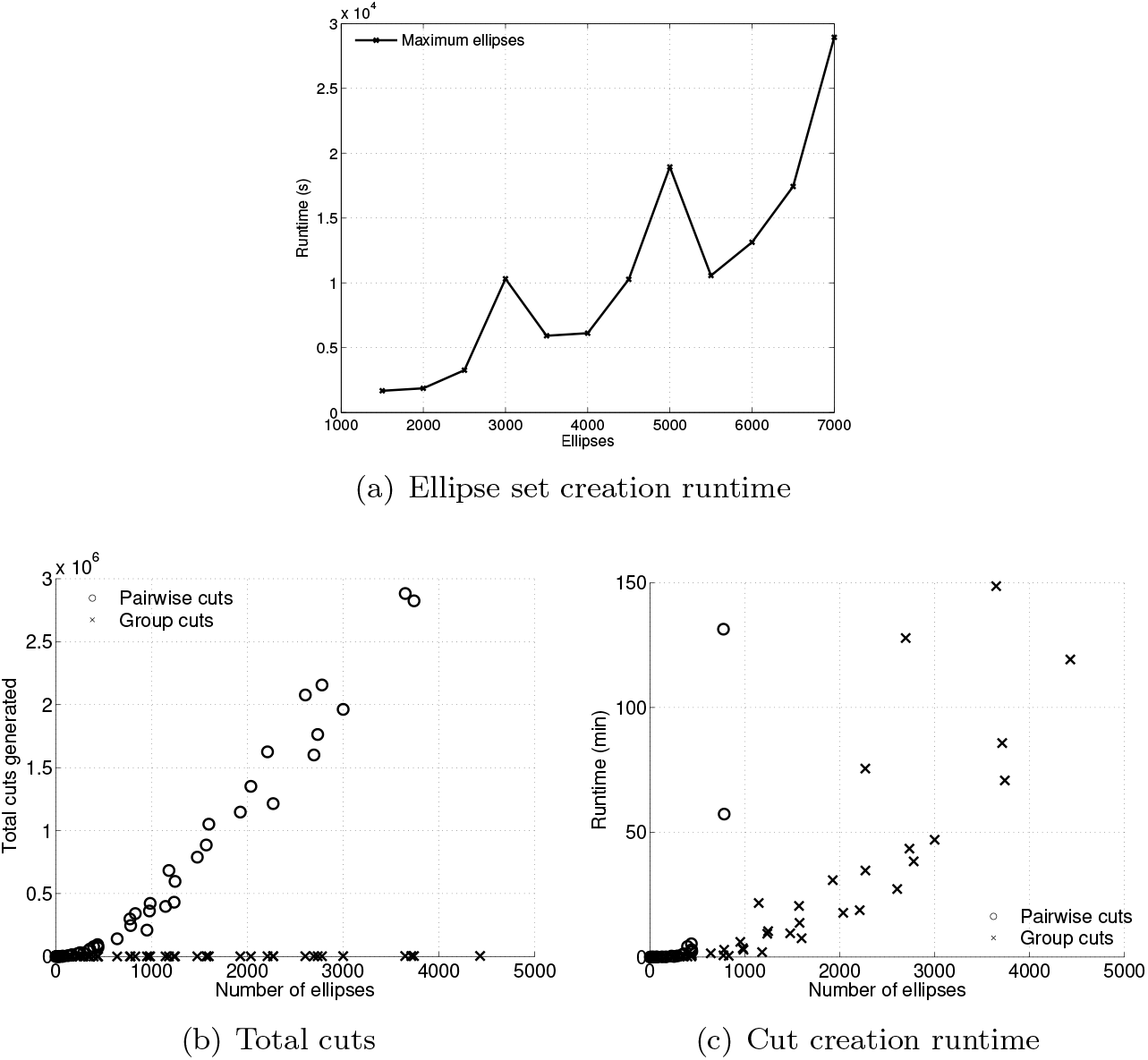
Ellipse and cut generation computational performance

The maximum number of pairwise no-good cuts and group cuts that can be generated is *m*!*/*(2 × (*m*− 2)!) and *m*, respectively. For our cases, the number of no-good cuts is up to 3,000,000, while only up to 4,500 group cuts are generated. As expected, the number of cuts generated and computation time required for pairwise cuts is significantly larger than for group cuts (Figure 2), and we therefore only present computational performance of RFA-SCP using group cuts (Figure 3). These cuts significantly improve the number of solvable instances compared to the full RFA-SCP model.

**Figure 3:**
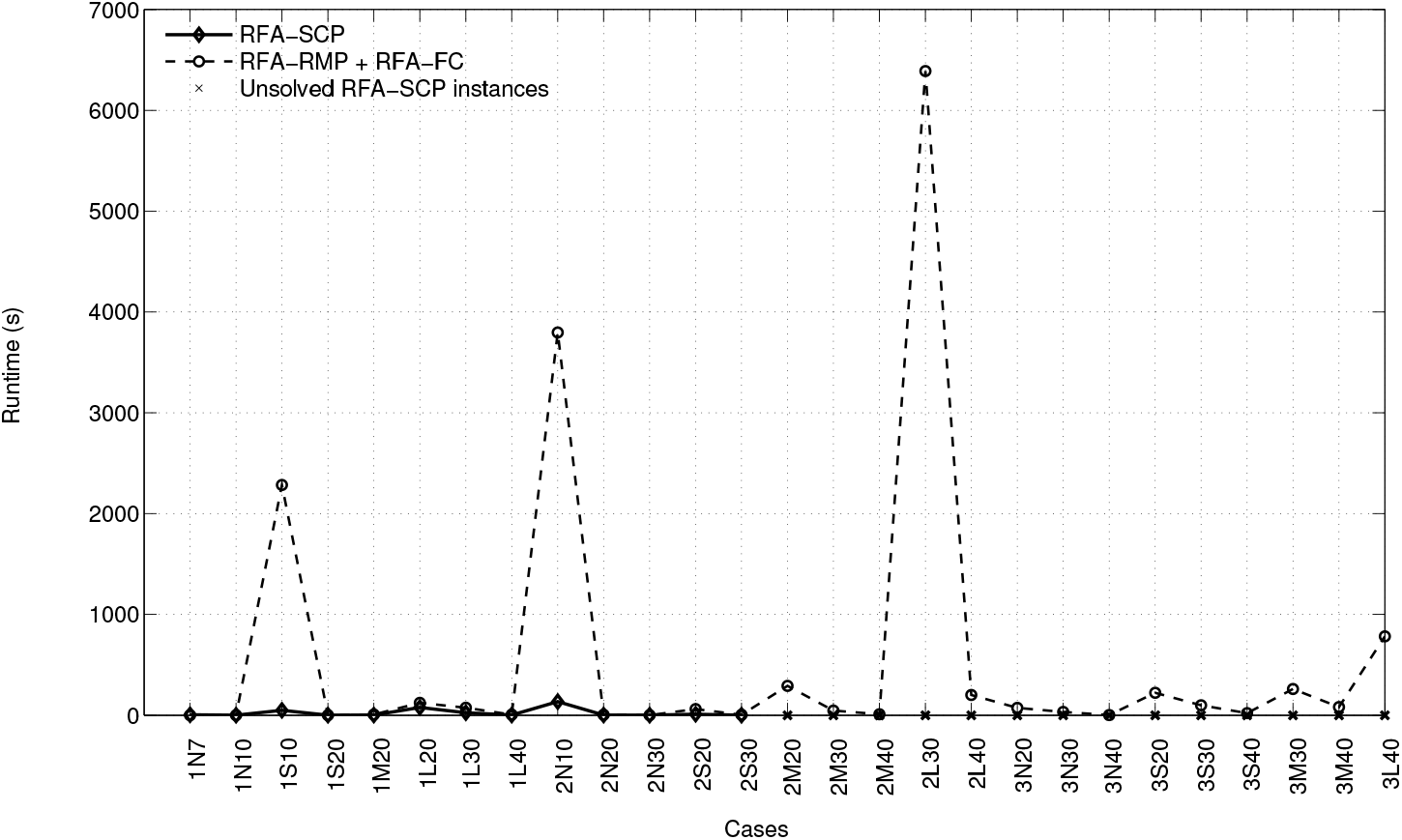
Computation times of RFA-SCP with row generation using group cuts and 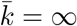

Total ablations and model feasibility depend primarily on the definition of ellipse sets (Table 3). When 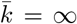, the solution provides a lower bound (*l*) on the minimum ablations required for full target coverage. When *k* ablations are desired, the models may be infeasible due to no pairwise valid needle positions or incomplete target coverage. For instance, for Case 1N, *ℓ* = 1 ablations are required for *λ* ≥7 and for *k* = 2 *> ℓ*, the model is infeasible due to intersecting needle placements. For Case 2S, *ℓ* = 4 ablations are required for *λ* = 10 and hence when *k* ∈{1, 2} *< ℓ*, the model becomes infeasible due to insufficient target coverage.

**Table 3:** Total ablations with 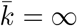 (RMP-B)

| Case ID | Tip length $\lambda$ mm | | | | |
| --- | --- | --- | --- | --- | --- |
|  | 7 | 10 | 20 | 30 | 40 |
| 1N | 1 | 1 | - | - | - |
| 1S | *inf* | 2 | 1 | - | - |
| 1M | *inf* | 3 | 1 | - | - |
| 1L | *inf* | *inf* | 1 | 1 | - |
| 2N | 4 | 3 | 1 | 1 | - |
| 2S | *inf* | 4 | 1 | 1 | - |
| 2M | *inf* | 7 | 1 | 1 | - |
| 2L | *inf* | *inf* | 2 | 1 | 1 |
| 3N | *inf* | 6 | 2 | 1 | 1 |
| 3S | *inf* | 9 | 2 | 1 | 1 |
| 3M | *mem* | 0 | 3 | 2 | 2 |
| 3L | *mem* | 0 | 4 | 2 | 2 |

The row generation on the target coverage (RMP-E + FC-E) performs 67% faster then RFA-SCP on more than 60% of our cases (Figure 4(a)); while, as seen in Figure 4(b), the model RFA-SCP outperforms row generation on pairwise validity matrix (RMP-B + FC-B). The runtime of the model is influenced by the target size as well as number of ablations desired. RMP-B + FC-B performs poorly because the RMP is resolved at each iteration with new cuts when more than a single ablation is required. However, RMP-E + FC-E outperforms RFA-SCP and RMP-B + FC-B because of up to 90% reduction in target coverage constraints (**E**), and in a multi-needle ablation scenario, the possibility of invalid needle placements is higher than the failure to cover internal target voxels due to a large number of needle combinations. Finally, although using 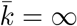 does not require a priori knowledge of the number of ablations, faster runtimes (up to *<* 50 min) can be achieved by bounding 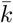, especially to detect infeasible or undesirable solutions (e.g., large number of ablations), with row generation on target coverage outperforming the full model (Figure 5).

**Figure 4:**
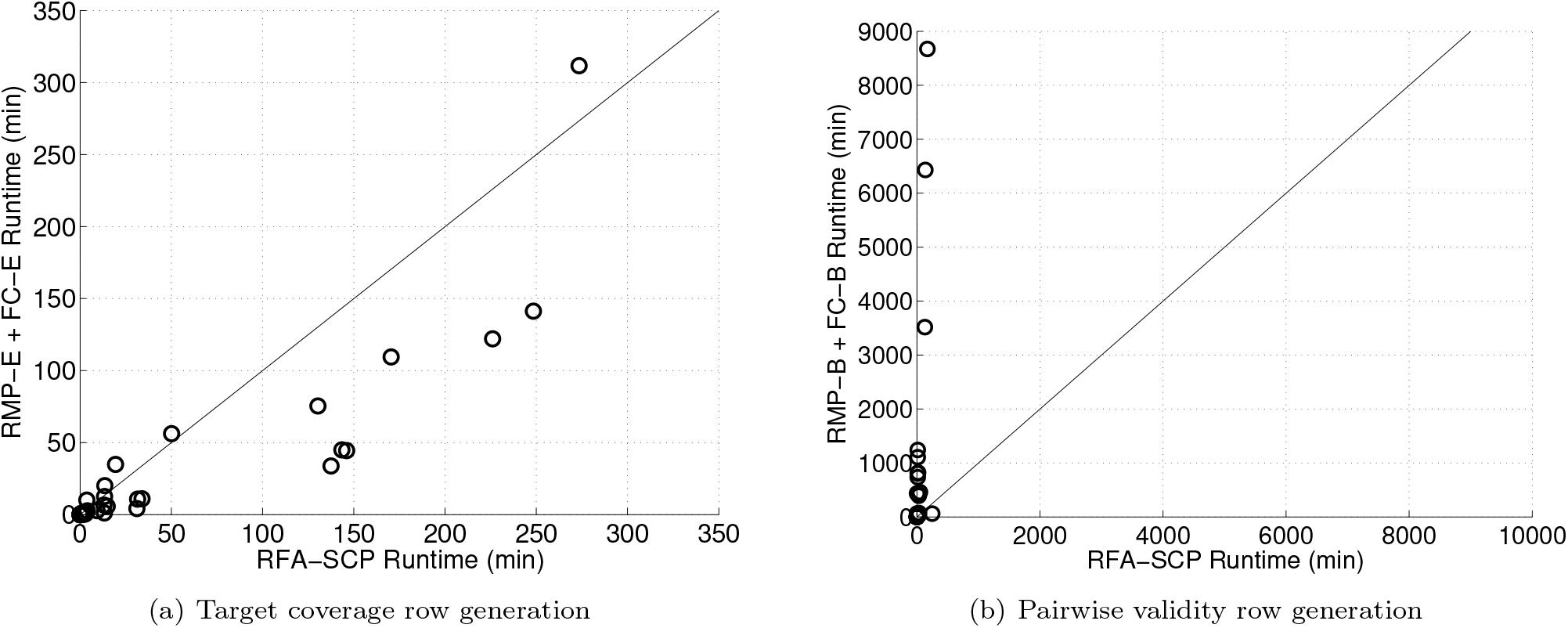
Computation times of row generation approaches v. full RFA-SCP model

**Figure 5:**
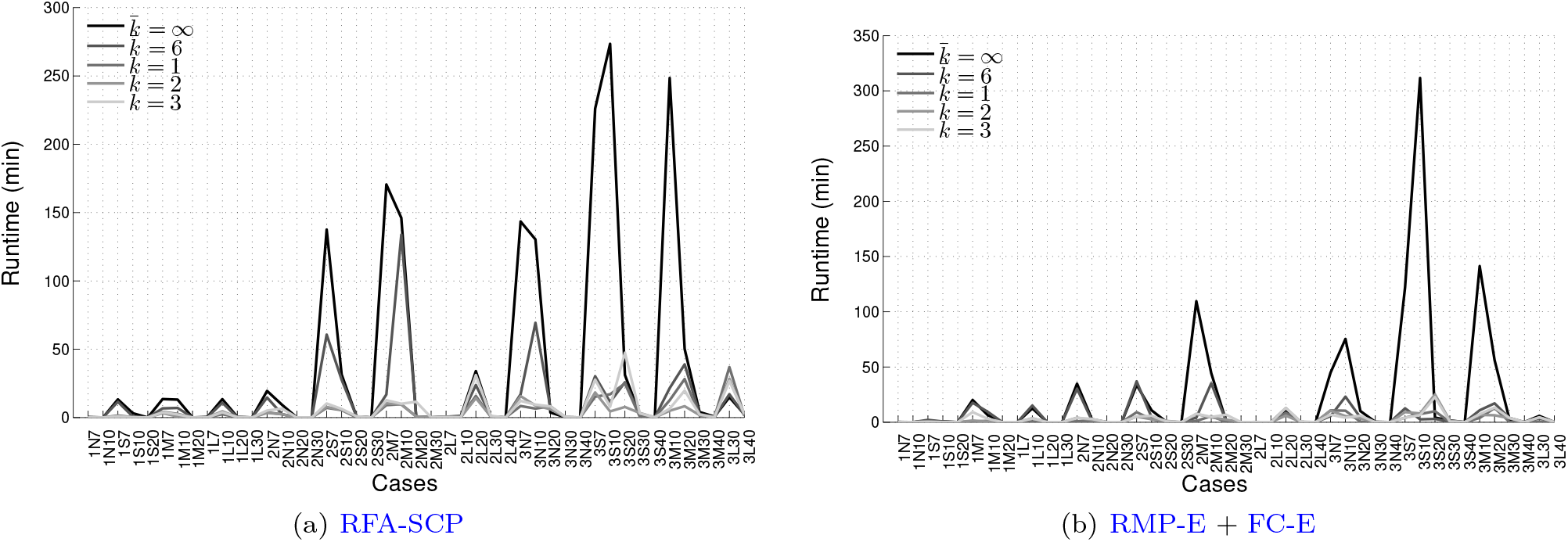
Computation times with bounded 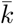

After obtaining needle trajectories (examples shown in Figure 6), we then determine the thermal dose distribution by TDO. We compute the BHTE, ATDM, and percentage damage for each target and needle length combination against 12 voltage values from 2.5 to 30 V in increments of 2.5V. The simulation time is 20 minutes and *dt* = 0.5 s. The average runtimes for Case 1 appear in Figure 7. Larger needle length causes higher lesion volumes (Figure 8), consequently higher target (Figure 9) and OAR damage volumes (Figure 10). While BHTE damage target coverage occurs at lower voltage values, ATDM and percentage damage models require higher voltage values (Figure 9).

**Figure 6:**
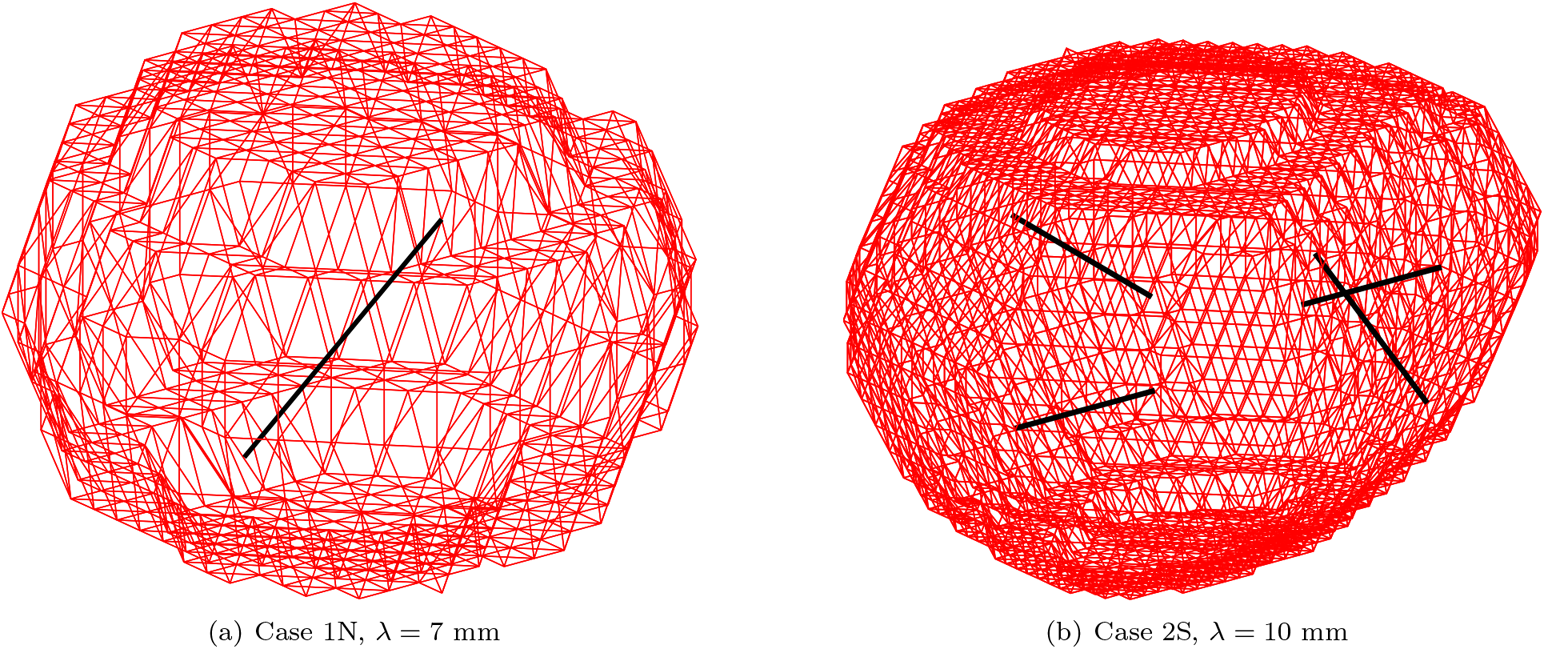
Needle placement for single and multiple needle ablation using trajectory planning

**Figure 7:**
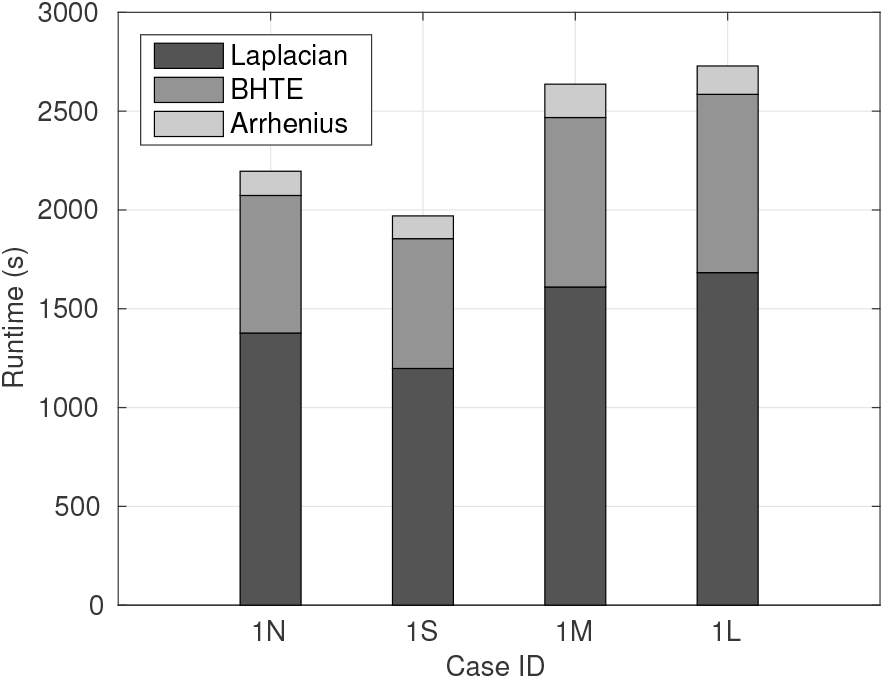
Average BHTE runtimes

**Figure 8:**
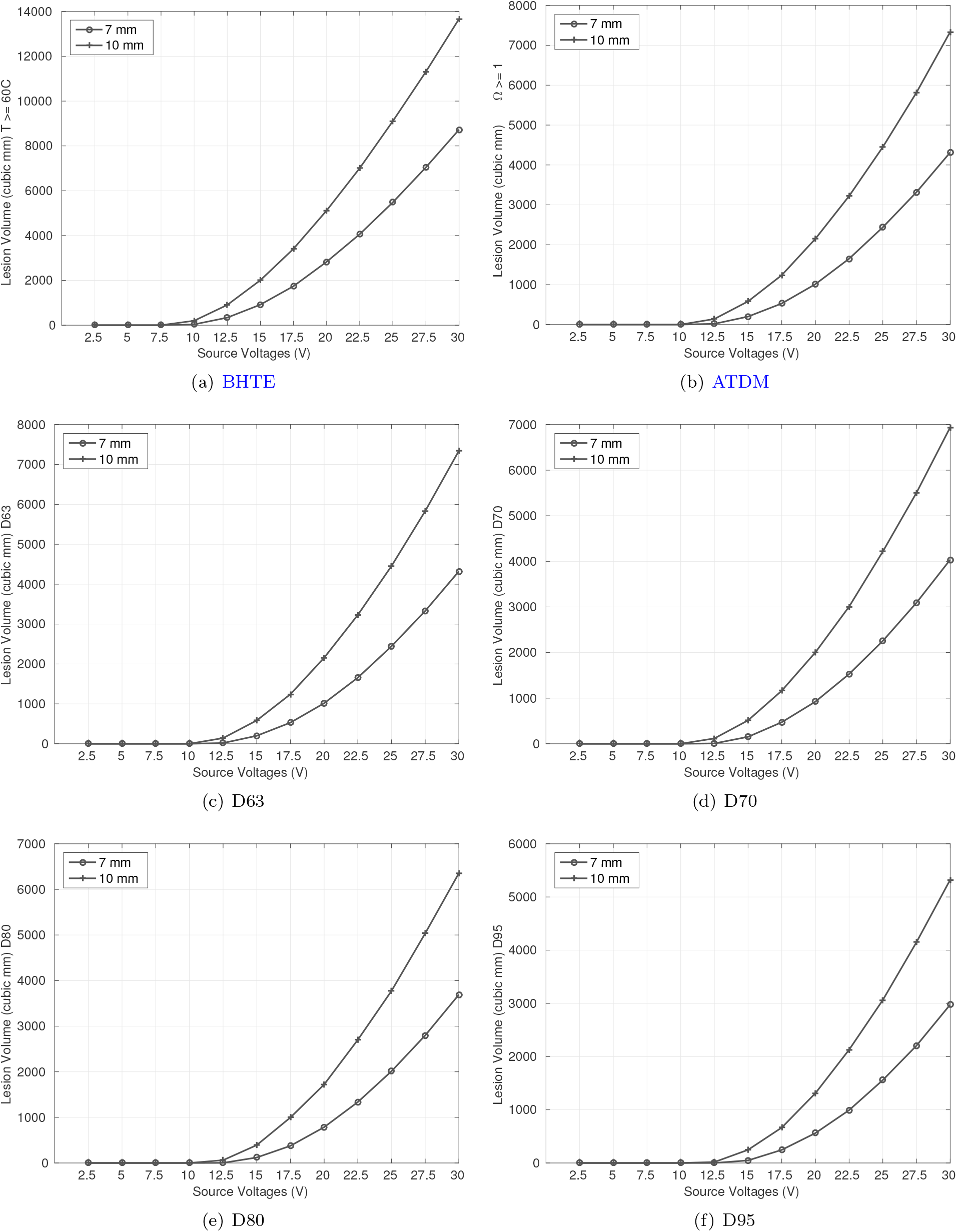
Lesion volumes (Case 1N)

**Figure 9:**
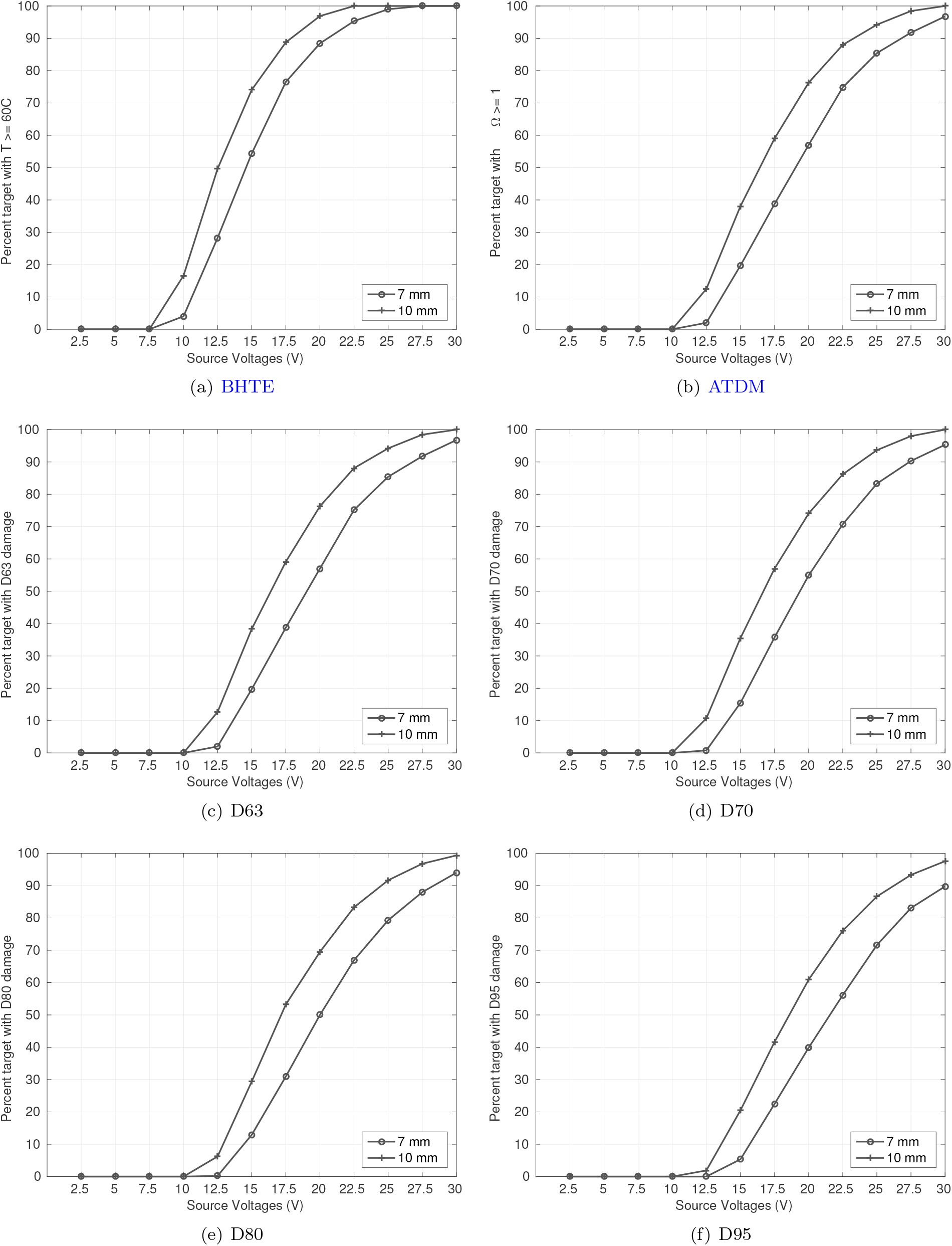
Percent target coverage (Case 1N)

**Figure 10:**
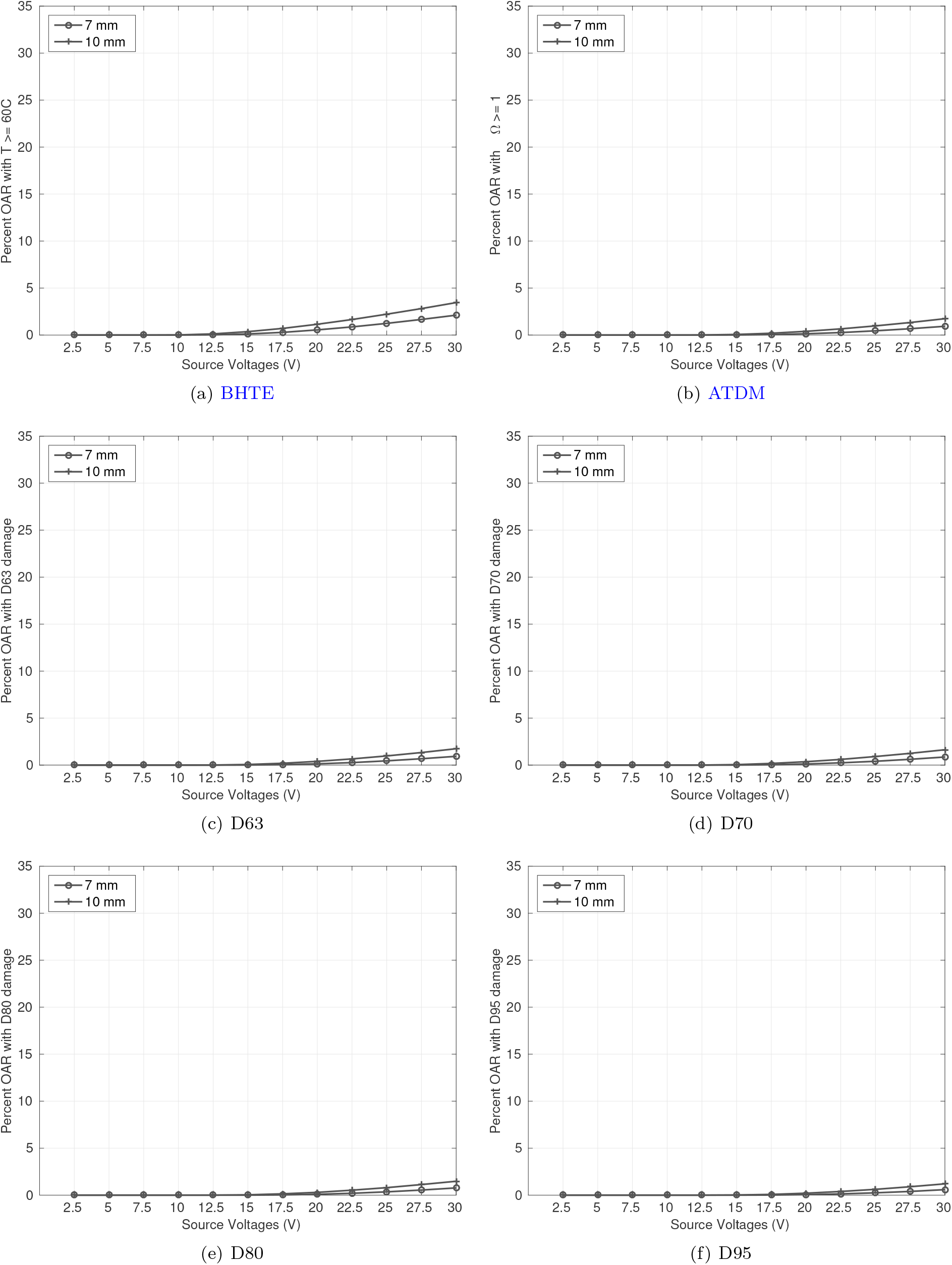
Percent OAR coverage (Case 1N)

## 5 Discussion

We extend our two-stage RFA planning framework for single, clustered, and multiple needle placement by incorporating trajectory planning [15] and for the first time present a full RFA planning framework that incorporates trajectory planing as well as thermal dose computations. In our previous work, we used mathematically rigorous convex optimization models for single and clustered needle placement. For multiple needle placement we used K-means to identify clusters, where each cluster corresponds to a single ablation needle, and used convex models to identify needle position and orientation for each cluster. Thus, the needle position corresponded to the geometric centroid of the target (or cluster) and the orientation to the their shape. In this work, we use IP models, solved to global optimality, to identify best needle positions for single or multiple needle placement. Thus, needle positions and orientations are discretized and therefore do not correspond to targets’ geometric centroid or shape. We are still able to attain full target coverage albeit at higher source voltage. Further, trajectory planning disregards large needle types for smaller target unlike our previous work where all needle lengths were explored for a target. Thus, our trajectory planning model provides a realistic advantage to our previous models.

Most RFA treatment planning systems that incorporate trajectory planning focus on single needle placement [12, 16, 17, 19], sequential integer programming techniques for multiple needle placement that result in suboptimal solutions [21, 22], and do not incorporate thermal dose computations. Existing simultaneous optimization models for RFA do not incorporate trajectory planning [11, 14] and are restricted to either single or clustered needle placement while k-means approach is used for multiple needle placement. Due to non-linearity of these ablation models, only a locally optimal solution is viable. Further, due to a PDE-constrained system, the models are restricted to ablation modality. Finally, unlike radiation planning systems there is a lack of standard based on either conformity or OAR-sparing to quantify treatment plans in ablation.

This work assumes an ablation radii for each individual needle inserted and approximates it to an ellipse. However, vendor specifications provide ablation radii for multiple needles where needles are placed parallely and operated simultaneously on a porcine liver. The shape of lesion is unclear when multiple needles are placed non-parallely due to lack of clinical experiments. Thus, the NOO stage may incorrectly estimate the ablation radius and consequently the target coverage. The target coverage can be determined by thermal dose simulations as well as by enforcing minimum number of ablations based on clinical experience. The algorithm for ellipse predefinition presented in this work can be enhanced by incorporating other criteria like path length as well as by visual representation of the needles. Similarly, the objective function of our SCP models can be modified to include a weighted sum using criteria like unablated target, path length, proximity to critical structures like veins, and total ablations. Finally, we recommend the use of commercial PDE solvers to enhance the quality of treatment.

## 6 Conclusion

We have presented a comprehensive framework to develop RFA treatment plans for liver cancer. For the first time we have incorporated trajectory planning as well as thermal dose simulations that considers critical structures before simultaneous multiple needle placement. Further, we propose a novel IP approach that uses row generation techniques, tractable to large cases, for needle placement. We also present algorithms for predefining needle paths that consider physical as well as clinical criteria. We solve these models using varying inputs and show promising results that achieve full target coverage. Before clinical applicability we propose testing the methodology with actual risk structures and thereby implementing better algorithm for ellipse predefinition.

## Data Availability

Data not available due to patient privacy

